# Quantitative insights into effects of intrapartum antibiotics and birth mode on infant gut microbiota in relation to well-being during the first year of life

**DOI:** 10.1101/2021.11.01.21265735

**Authors:** Katri Korpela, Roosa Jokela, Ching Jian, Evgenia Dikareva, Anne Nikkonen, Terhi Saisto, Kirsi Skogberg, Willem M. de Vos, Kaija-Leena Kolho, Anne Salonen

## Abstract

**Background and aims:** Caesarean section (CS)-birth and maternally administered intrapartum antibiotics (IP) affect colonization of the neonate. We compared the effects of CS delivery and IP antibiotics on infant gut microbiota development and wellbeing over the first year. To understand the developing community dynamics, we focused on absolute bacterial abundance estimates over relative abundances.

**Methods:** We studied 144 healthy infants born between gestational weeks 37-42 vaginally without antibiotics (N=58), with IP penicillin (N=25) or cephalosporin (N=13), or by CS with IP cephalosporin (N=34) or other antibiotics (N=14). Gut microbiota composition and temporal development was analysed at 5-7 time points during the first year of life using 16S rRNA gene amplicon sequencing, complemented with qPCR to obtain absolute abundance estimates in 92 infants. A mediation analysis was carried out to identify taxa linked to gastrointestinal function and discomfort (crying, defecation frequency and signs of gastrointestinal symptoms) and birth interventions.

**Results:** Based on absolute abundance estimates, depletion of *Bacteroides* spp. was specific to CS birth while decreased bifidobacteria and increased Bacilli were common to CS birth and exposure to IP antibiotics in vaginal delivery. Abundance of numerous taxa differed between the birth modes among cephalosporin-exposed infants. Penicillin had a milder impact on the infant gut microbiota than cephalosporin. The effects of both CS birth and IP antibiotics on infant gut microbiota associated with increased gastrointestinal symptoms during the first months.

**Conclusion:** CS birth and maternal IP antibiotics have both specific and overlapping effects on infant gut microbiota development. The resulting microbiota deviations were found to associate with gastrointestinal symptoms in infancy.

**What You Need to Know:** *Background and Context:* Birth mode and maternal intrapartum antibiotics affect infant’s gut microbiota development but their relative contribution, and effects on absolute bacterial abundances and infant health remain unknown.

*New Findings:* Utilizing quantitative microbiota profiling, we identified shared and unique microbiota effects of birth mode and intrapartum antibiotics which explained up to 54% of variation in parent-reported gastrointestinal symptoms in infants.

*Limitations:* Due to the limited sample sizes, especially during the first weeks of life, stratified analyses according to antibiotic dosing could not be performed, and the results on gastrointestinal symptom-microbiota-associations are tentative.

*Impact:* Birth mode overrules the effects of maternal antibiotics on infant microbiota development, while both birth mode and maternal antibiotic use are associated to common functional gastrointestinal symptoms in infancy.

*Lay summary:* Caesarean-section birth and maternal antibiotics during vaginal birth affect infant’s gut microbiota and may increase gastrointestinal discomfort.

## Introduction

The human intestinal microbiota develops via successional stages during early life.^1^ The infant’s gut microbiota contributes to early immunological^2^ and metabolic programming^3^ that affects host health later in life.^4^ Transmission of maternal microbes to the offspring during birth plays a major role in the initial colonization.^1, 5, 6^ The mode of birth is a major determinant of the neonates’ initial microbial exposure; the assembly and dynamics of intestinal microbiota in infants born via caesarean section (CS) versus vaginal delivery (VD) differ substantially,^1, 7^ which has been suggested to mediate the negative long-term health outcomes related to CS birth.^8, 9^

CS birth is not the only intervention impacting the neonate’s early colonisation. Intrapartum (IP) antibiotic prophylaxis is routinely administered to mothers undergoing CS to reduce the risk of post-caesarean maternal infection. In vaginal deliveries approximately 25% of the mothers receive antibiotics to prevent neonatal group B Streptococcus (GBS) infection.^10^ Thus, antibiotics are used in a substantial number of deliveries in developed countries. Emerging evidence suggests that IP antibiotics influence the infant’s microbiota development over a period of several months.^11-13^

Previous studies utilizing next generation sequencing (NGS) have characterised the infant gut microbiota using relative bacterial abundance. However, quantitative analyses indicate that early microbiota development entails major quantitative changes both for the total bacterial load as well as on individual taxa,^14^ and the effects of birth interventions on absolute bacterial profiles are unknown.^15^ In addition, current studies addressing the health effects of IP on newborns have largely focused on neonatal GBS infection,^16, 17^ with first reports investigating their potential long-term effects on child health.^18^ We analysed the gut microbiota development in 144 healthy infants over the first year of life, focusing on 92 infants from whom qPCR- and NGS-based quantitative microbiota profiling^19^ data for absolute abundance estimation was available. The bacterial abundances were further associated with parallelly collected gastrointestinal (GI) and general health data. This approach enabled us to bypass the compositionality problem arising from relative abundances,^20^ which may mask true community dynamics^21^ and lead to high false discovery rates.^19, 22^ We controlled for potential confounding factors using a carefully selected cohort, aiming to disentangle the effect of CS birth and IP antibiotics from potentially collinear effects.

## Materials and Methods

### Study design

Infants specifically recruited for this study (Jorvi cohort, N=68) and additional 83 infants recruited for the Finnish Health and Early Life Microbiota (HELMi) cohort^23^ (<u>NCT03996304</u>) were included in the study. In both cases, pregnant women with singleton gestation were recruited from the general population in Southern Finland. Healthy babies, born on gestational weeks 37-42, without known congenital defects and exceeding the birth weight of 2.5 kg were included in the study (except one baby with birth weight of 2.4kg). At least one parent had to be Finnish speaking to be able to answer the questionnaires.

For the Jorvi cohort, 31 mothers delivered with (vaginal deliveries N=26/31) and 37 without IP antibiotics (all vaginal deliveries). Infant faecal samples were collected on day 1, 2, and 7, and at 1, 3, 6 and 12 months. From the HELMi cohort, 83 infants were selected based on birth mode, IP antibiotics, and the availability of sequencing data from infant’s faecal samples at the age of 3 (considered as 1 month sample), and 6 weeks, and 3, 6, 9, and 12 months. From the combined cohort, five infants were excluded as they received post-natal antibiotics prior to the first sampling point, and two infants were excluded for low sequence quality, leaving a total of 144 infants. Additionally, all samples following a post-natal antibiotic treatment were excluded from the analysis as well as individual samples with low sequence quality (see Statistical analysis). We divided the remaining infants into five study groups based on birth mode and IP antibiotics: vaginally delivered (VD) reference group without IP antibiotics (N=58), and four exposure groups: cefuroxime or cephalexin (cep) in CS (N=34), or in VD (N=13), penicillin (pen) in VD (N=25), or in CS penicillin or any other antibiotics, a combination of antibiotics or the antibiotic class not reported (CS-other, N=14) (Table 1). The CS groups comprised elective (N=23) and urgent (N=25) cases; all involved IP antibiotics. Based on sample sufficiency, birth mode and sampling point the samples from 92 infants were quantified for total bacteria by qPCR,^19^ representing 26 infants in the reference group VD, 7 in VD-cep, 13 in VD pen, 33 in CS-cep, and 13 in CS-other group.

**Table 1.** Faecal samples analysed per delivery group and age (relative data /absolute data).

|  | VD | VD-cep | VD-pen | CS-cep | CS-other |
| --- | --- | --- | --- | --- | --- |
| 1 day | 11/0 | 1/0 | 4/0 | 2/0 | 0/0 |
| 2 days | 23/4 | 2/0 | 7/5 | 5/2 | 2/1 |
| 1 week | 31/1 | 3/0 | 13/0 | 7/0 | 0/0 |
| 1 month | 46/25 | 12/7 | 23/12 | 33/32 | 13/13 |
| 6 weeks | 20/20 | 10/7 | 10/8 | 25/25 | 12/12 |
| 3 months | 41/20 | 12/7 | 22/8 | 31/27 | 13/12 |
| 6 months | 48/22 | 12/6 | 24/12 | 32/30 | 13/12 |
| 9 months | 16/16 | 7/4 | 9/7 | 20/20 | 10/8 |
| 12 months | 36/3 | 8/0 | 15/3 | 12/4 | 3/1 |

VD = vaginal delivery, cep = cephalosporin, pen = penicillin, CS = Caesarean section

Both cohort studies were approved by the ethical committee of The Hospital District of Helsinki and Uusimaa and performed in accordance with the principles of the Helsinki Declaration. Parents signed an informed consent at enrolment.

### Samples and data collection

Parents collected faecal samples from the infants and stored them at -20°C. The samples were transported frozen to the study centre within 6 months for -80°C storage. Data on feeding regime, probiotic and antibiotic use, child health and growth were collected with questionnaires (HELMi) or via phone interviews from the parents at the times of stool sampling (Jorvi). Data on exposures *i*.*e*., the mode of delivery and the usage, type and dosing of maternal IP antibiotics were collected from hospital records for both cohorts.

### Health outcomes

Infant health and wellbeing were addressed using questionnaire data available for the HELMi cohort. We used data from weekly to monthly repeating questionnaires using the questionnaire closest to each faecal sampling. The variables included were related to crying (hours per day and intensity estimated by a 0-100 mm visual analogue scale (VAS), collected until six months, GI function (defecation rate, parent-perceived stomach pain intensity and flatulence) until 9 months, and frequency of infections estimated by the number of post-natal antibiotic courses by age of 12 months. Infant growth was measured in postnatal care visits and transformed to age-dependent WHO z-scores.^24^

### Microbiota profiling

Bacterial DNA was extracted from the faecal samples using a previously described bead beating method^25^ and KingFisherTM Flex automated purification system (ThermoFisher Scientific, USA) as detailed in Supplementary methods. 16S rRNA gene amplicon sequencing was performed using Illumina MiSeq and HiSeq platforms for V3-V4 or V3 region as previously described,^19, 26^ respectively. In case of low DNA yield or unsuccessful sequencing, the default library preparation protocol was modified as specified in Supplementary methods. The numeration of total bacteria was performed in triplicate as previously described.^19^ Bacterial DNA was quantified by amplifying 0.5 ng aliquots of each DNA extract with universal bacterial primers 331F/797R^27^ targeting the 16S rRNA gene. The standard curves ranging from 10^2^ to 10^7^ copies were constructed using the full-length amplicons of 16S rRNA gene of *Bifidobacterium bifidum* to convert the threshold cycle (Ct) values into 16S rRNA gene copy numbers per g of faeces.

The sequencing reads were processed using R package mare,^28^ which relies on USEARCH^29^ for quality filtering, chimera detection, and taxonomic annotation. Only the forward reads (V3), truncated to 150 bases, were used.^30^ Reads occurring <10-50 times, depending on run size, were excluded as potentially erroneous. Taxonomic annotation was performed using USEARCH^29^ by mapping the reads to the SILVA 16S rRNA reference database version 115,^31^ restricted to gut-associated taxa. Further, potential contaminants in the low-DNA-yield samples were filtered by removing reads appearing in negative controls (PCR or extraction blanks) in corresponding numbers from all samples. The absolute abundances were estimated and 16S rRNA gene copy-number corrected as previously described.^19^

### Statistical analysis

After processing, the median number of reads obtained was 26 384 (range 3 to 253 458). Sufficient sequencing coverage was evaluated by comparing observed species richness to read count, and a cut-off of 2000 reads was chosen for HELMi samples, and 120 for Jorvi samples (to accommodate low-biomass samples from the first week of life). Samples with insufficient coverage (N=138, largely samples from the first week of life) and 69 samples with extremely high read counts from a single HiSeq run with high proportions of probable contaminants (mostly *Alcaligenaceae*)^32^ were excluded, leaving a total of 688 high-quality samples from 144 infants for downstream analyses, and 393 high-quality qPCR quantified samples from 92 infants.

The statistical analysis was conducted in R version 3.6.3 with the package *mare*,^28^ with tools from packages *vegan*,^33^ *MASS*,^34^ and *nlme*^35^, detailed in Supplementary methods. The association of antibiotic exposure (no antibiotic, penicillin, cephalosporin) and birth mode (vaginal or CS) with health outcomes was analysed at each faecal sampling until 9 months using a negative binomial model adjusting for feeding type and probiotic use. To determine if the differences in health outcomes were mediated by changes in gut microbiota, the *PathModel* function in *mare*^28^ was used to identify taxa predicting symptoms and the role of birth interventions related to the predictive taxa.

## Results

### Study population

An average of 4.9 samples (range 1-7) was collected per infant (Table 1) from the 144 infants born by VD (N=96) or CS (N=48) during the first12 months. The urgent and elective CSs were pooled for analysis (Supplementary results, Supplementary Figure 1). The IP antibiotics and doses varied among the delivery modes, with cefuroxime being the most common antibiotic used in CS and penicillin in VD (Table 2). Of all the VD antibiotics, 74% were solely prophylactic, of which 93% to prevent GBS transmission, and 7% due to premature rupture of membranes, while 69% of cephalosporins were therapeutic. Therapeutic IP antibiotics were administered due to fever or a rise in C-reactive protein (CRP) of the mother. Absolute bacterial profiles were calculated for a subset of 92 infants (VD: N=46, CS: N=46). Infant background statistics are summarised in Supplementary Table 1.

**Table 2.** Intrapartum antibiotics and doses by delivery group among the 144 infants (relative data / absolute data).

| Birth mode | antibiotic | 1 dose | 2 or more doses | Total count of infants |
| --- | --- | --- | --- | --- |
| VD | penicillin | 4/1 | 21/12 | 25/13 |
|  | cefuroxime | 8/3 | 5/4 | 13/7 |
| Elective CS | cefuroxime | 13/12 |  | 13/12 |
|  | cephalexin | 7/7 |  | 7/7 |
|  | clindamycin | 1/1 |  | 1/1 |
|  | metronidazole | 1/1 |  | 1/1 |
|  | not reported | 1/1 |  | 1/1 |
| Urgent CS | cefuroxime | 10/10 | 1/1 | 11/11 |
|  | cephalexin | 3/3 |  | 3/3 |
|  | clindamycin | 2/2 | 2/2 | 4/4 |
|  | penicillin |  | 2/1 | 2/1 |
|  | combination | 1/1 | 4/4 | 5/5 |

VD = vaginal delivery, CS = Caesarean section.

### The early development of the gut microbiota from birth to 12 months of age

Birth mode or antibiotic exposure did not influence the total bacterial abundance quantified by qPCR at any given sampling time (*P*>0.05). Overall, the bacterial load increased with infant age in all groups, especially during the first weeks (Supplementary Figure 2).

To understand the temporal dynamics of the gut microbiota in infants, we analysed the average developmental trajectory of the 6 most abundant bacterial classes in infants born by VD without antibiotics, comparing their relative (day 1 onwards) and absolute abundances (day 2 onwards) (Figure 1). Relative abundances show different community dynamics, such as a gradual decrease in the relative abundance of *Enterobacteriales* instead of the peak in absolute abundance around 20 weeks of age (Figure 1).

**Figure 1.**
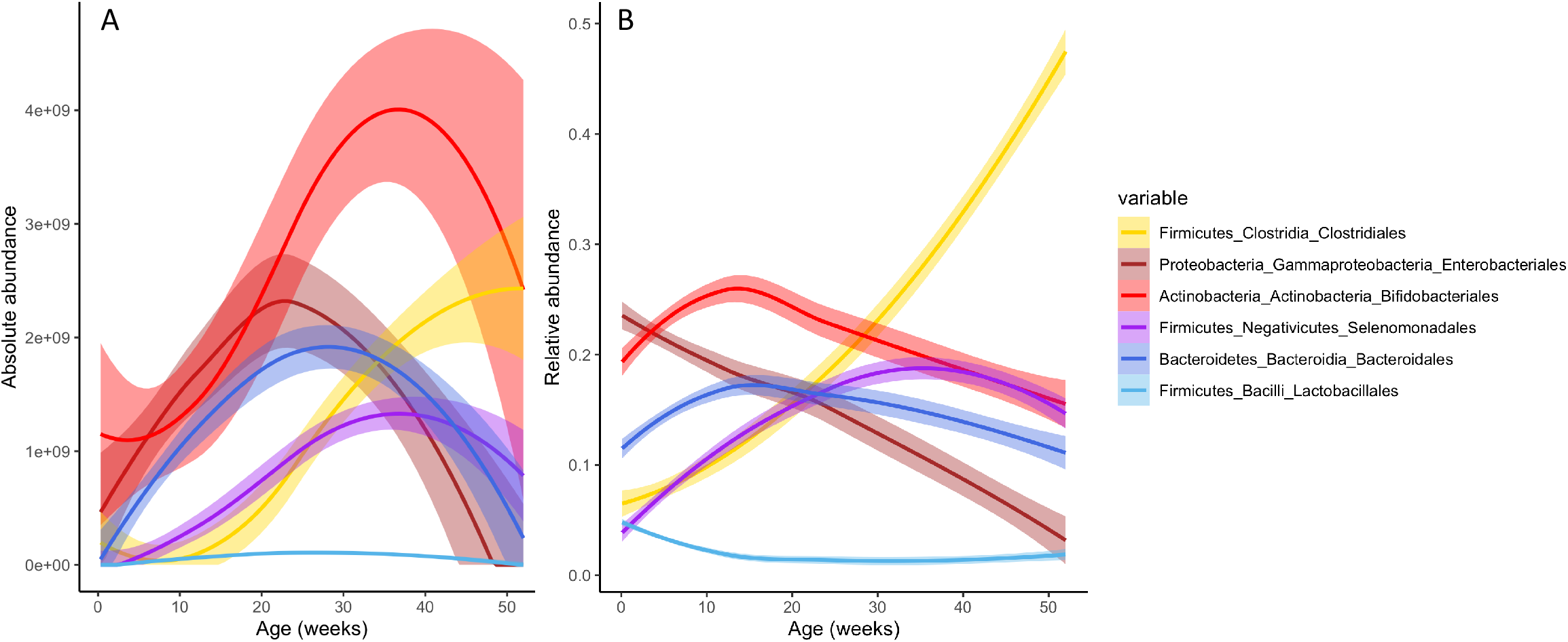
Temporal development of six most abundant bacterial classes in absolute (A) and relative (B) abundance in vaginally born infants not exposed to antibiotics. The lines show means with 50% confidence interval.

The overall developmental trajectories of the microbiota differed between the study groups categorized by delivery mode and antibiotic type (Table 1) in the ordination space using principal coordinate analysis (PCoA) (Figure 2). The temporal development of the gut microbiota followed a discernible pattern in the reference group (VD without antibiotics), and each exposure group showed a distinct developmental trajectory based on both absolute (Figure 2A) and relative data (Figure 2B). By 24 weeks, all but VD-cep converged with the reference group. The temporal trajectory of the gut microbiota in the VD-cep group markedly deviated from the reference group (Figure 2).

**Figure 2.**
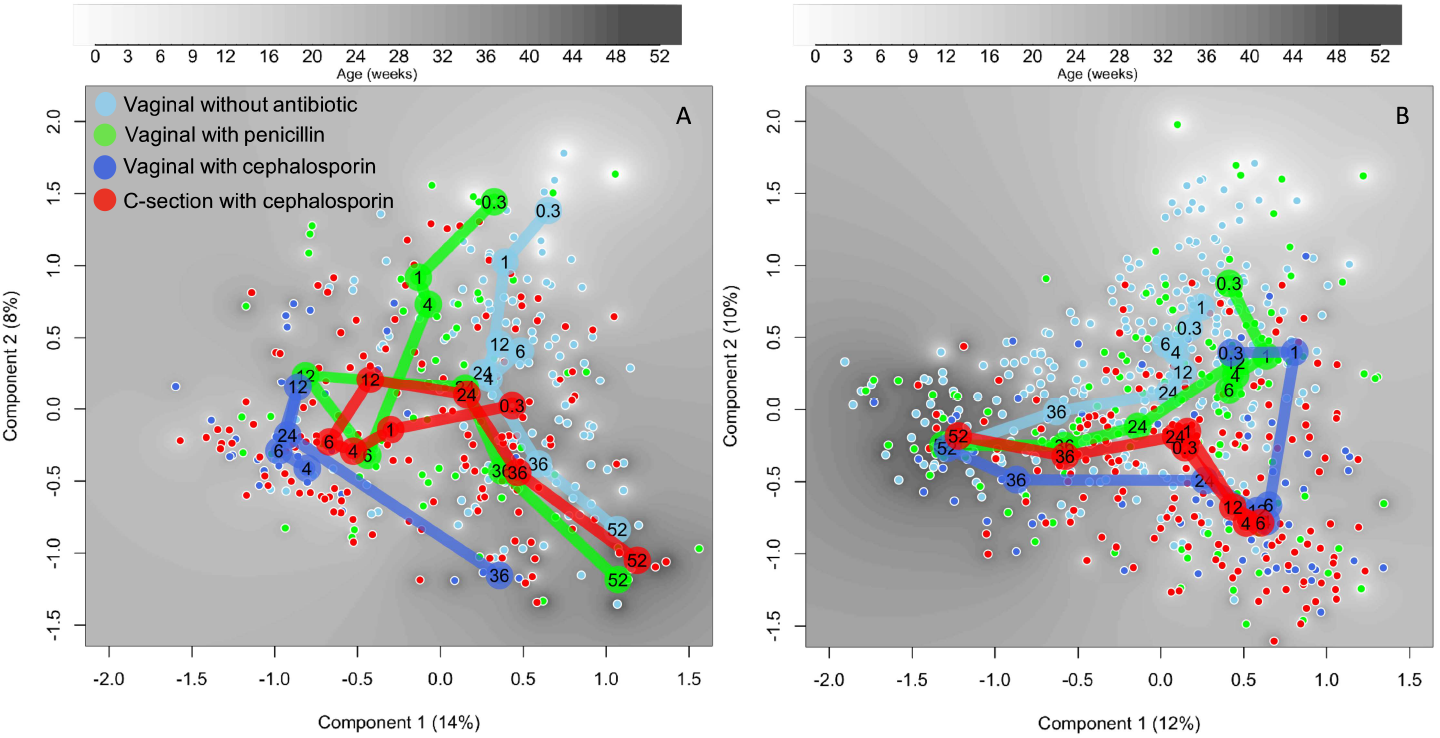
Principal coordinates analysis of the species-level absolute (A) and relative (B) abundances. Distances calculated based on Pearson correlations. The samples are coloured by treatment group and the mean component scores of the groups by age (weeks) presented by large circles.

### Impact of birth mode and intrapartum antibiotic on the temporal development of the gut microbiota using absolute data

To better understand the developing community dynamics, from this onwards we focused on the absolute abundance data. We first compared abundances of bacterial families and genera of the exposure groups to the reference (VD without antibiotic), adjusting for breastfeeding, probiotics, and the introduction of solids when relevant. The results observed at family level (Figure 3) were largely consistent at the genus level (Supplementary Figure 3).

**Figure 3.**
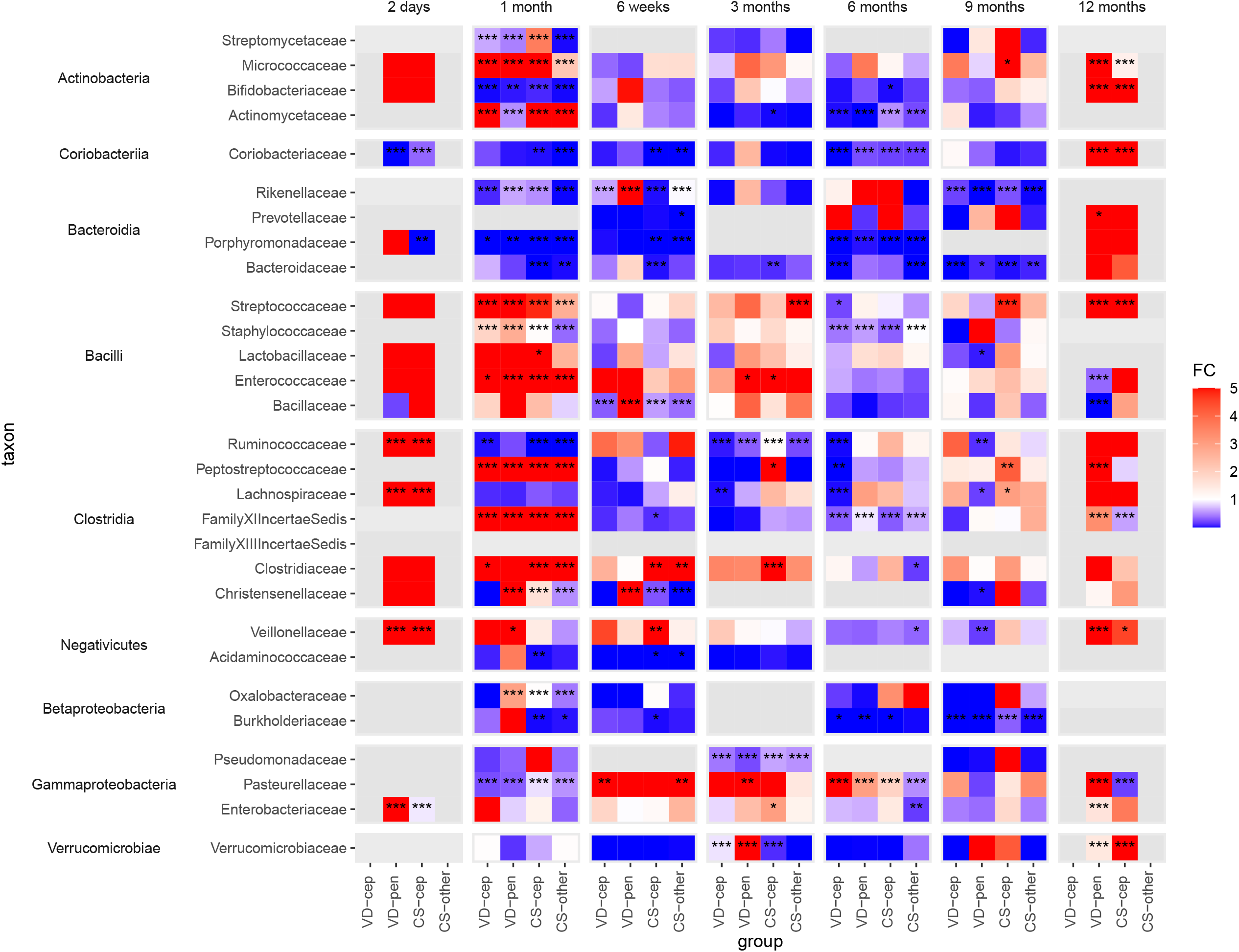
Fold changes calculated using absolute abundances of bacterial families with at least one group significantly differing from the reference group (VD) in the taxon-wise comparisions between all delivery groups. VD: vaginal delivery without antibiotic, VD-cep: vaginal delivery with cephalosporin, VD-pen: vaginal delivery with penicillin, CS-cep: C-section with cephalosporin, CS-other: C-section with any other antibiotic; FC: fold change (cut-off at 5 and 0.001 for clarity); *: *P*<0.05, **: *P*<0.01, ***: *P*<0.001, in all cases FDR<0.1.

In the first 3 months, the absolute abundance of *Bacteroidaceae* was significantly reduced in the CS, but not in the antibiotic-VD groups (fold change range 0.010-0.11, *P*<0.01 Figure 3). The depletion of *Bacteroidaceae* became statistically significant later, at 6 months and 9 months, in VD-cep (fold change 0.014, *P*=0.0003) and VD-pen (fold change 0.030, *P*=0.01).

*Coriobacteriaceae* (Actinobacteria) followed a pattern similar to that of *Bacteroidaceae*, being mainly associated with birth mode. The absolute abundance of *Bifidobacteriaceae* was strongly and negatively affected by both CS birth and by both cephalosporin and penicillin at 1 month, (fold change range 0.0040-0.18, *P*<0.01, Figure 3) compared to VD without antibiotics. *Actinomycetaceae* and *Micrococcaceae* were enriched in all the exposure groups except the VD-pen group at one month (fold change range 6.2-13, *P*<0.001, Figure 3). Families in the Bacilli class, *Staphylococcaceae, Enterococcaceae, Lactobacillaceae*, and *Streptococcaceae*, were increased in absolute abundance in all exposure groups during the first 3 months (fold change range 1.0-240, *P*<0.05, Figure 3). The absolute abundances of several families in the classes Clostridia and Negativicutes were increased in all the exposure groups during the first month in comparison to the reference, with a few exceptions: reduced *Christensenellaceae* in the CS-other group at 1 month and in both CS groups at 6 weeks, reduced *Ruminococcaceae* in all but the VD-pen group at 1 and 3 months, and reduced *Acidaminococcaceae* in the CS groups at 4-6 weeks (Figure 3). The absolute abundances of Proteobacterial families in the antibiotic-exposed groups significantly deviated from the reference at multiple time points (fold change range 0.00054-6100, *P*<0.05, Figure 3).

### Dissecting the effects of birth mode and intrapartum antibiotic on absolute levels of infant microbiota

We compared the birth modes while controlling for the IP antibiotic exposure, including only the cephalosporin-exposed CS and VD, to understand the effect of birth mode independently of antibiotic treatment, and *vice versa* (including only the vaginally born) (Supplementary Figures 4, 5). Several bacterial families differed in absolute abundance between the VD-cep and CS-cep groups, including enrichment of *Verrucomicrobiaceae* (3.8-fold difference, *P*<0.001 at 6 weeks), *Bifidobacteriaceae* (18 and 12-fold differences, *P*<0.05 at 1 and 3 months, respectively), *Streptomycetaceae* (7.0 and 62-fold differences, *P*<0.001 at 1 month and 9 months), *Lactobacillaceae* (8.3 and 12 -fold differences, *P*<0.05 at 3 and 9 months), *Bacillaceae* (3.8 and 8.5-fold differences, *P*<0.001 at 6 weeks and 6 months), *Peptostreptococcaceae* (fold change range 7.5-5900, *P*<0.01 at 1, 3 and 6 months), *Lachnospiraceae* (20 and 99-fold differences, *P*<0.01 at 3 and 6 months), *Clostridiaceae* (23-fold difference, *P*<0.001 at 1 month), and *Christensenellaceae* (210-fold difference, *P*<0.001 at 6 months) in the CS-cep group. In contrast, *Bacteroidaceae* was reduced in CS-cep in comparison to VD-cep during months 1-3 (fold change 0.07, *P*<0.01 at 6 weeks).

In terms of the effect of antibiotic types in the vaginally born infants, increased absolute abundances were found in *Verrucomicrobiaceae* and *Rikenellaceae* (22 and 79-fold differences, *P*<0.001 at 6 weeks), *Bifidobacteriaceae* and *Lactobacillaceae* (25 and 5.2-fold differences, *P*<0.01 at 3 months), *Coriobacteriaceae* (fold change range 6.0-910, *P*<0.001 at 1-3 months), *Peptostreptococcaceae* (94 and 72-fold differences, *P*<0.05 at 1 and 6 months), *Christensenellaceae* (8300-fold difference, *P*<0.001 at 6 months), *Bacillaceae* (36 and 13-fold differences, *P*<0.001 at 6 weeks and 6 months) in the VD-cep group compared to VD-pen. Members of *Clostridiaceae* were consistently reduced in VD-pen in comparison to VD-cep at 1 month (fold change 0.34, *P*<0.001).

### Health outcomes

To determine whether the mode of delivery and IP antibiotics were associated with the infants’ health during the first year of life, we compared the study groups in terms of health variables from the HELMi questionnaires pertinent to parent-reported digestive and overall health (Figure 4). All exposure groups showed increased GI symptoms mainly during the first 3 months compared to the reference group VD. There were significant differences in defecation rate, crying intensity, duration of crying per day, parent-perceived stomach pain intensity, and flatulence between the study groups (Figure 4). Infants’ overall health as reported by the parents did not differ between the study groups at any sampling age (ANOVA: *P*>0.1), nor did the cumulative number of antibiotic courses by 12 months (*P*=0.81) nor infant weights at birth or at 12 months (ANOVA, P≥0.25, Supplementary Table 1).

**Figure 4.**
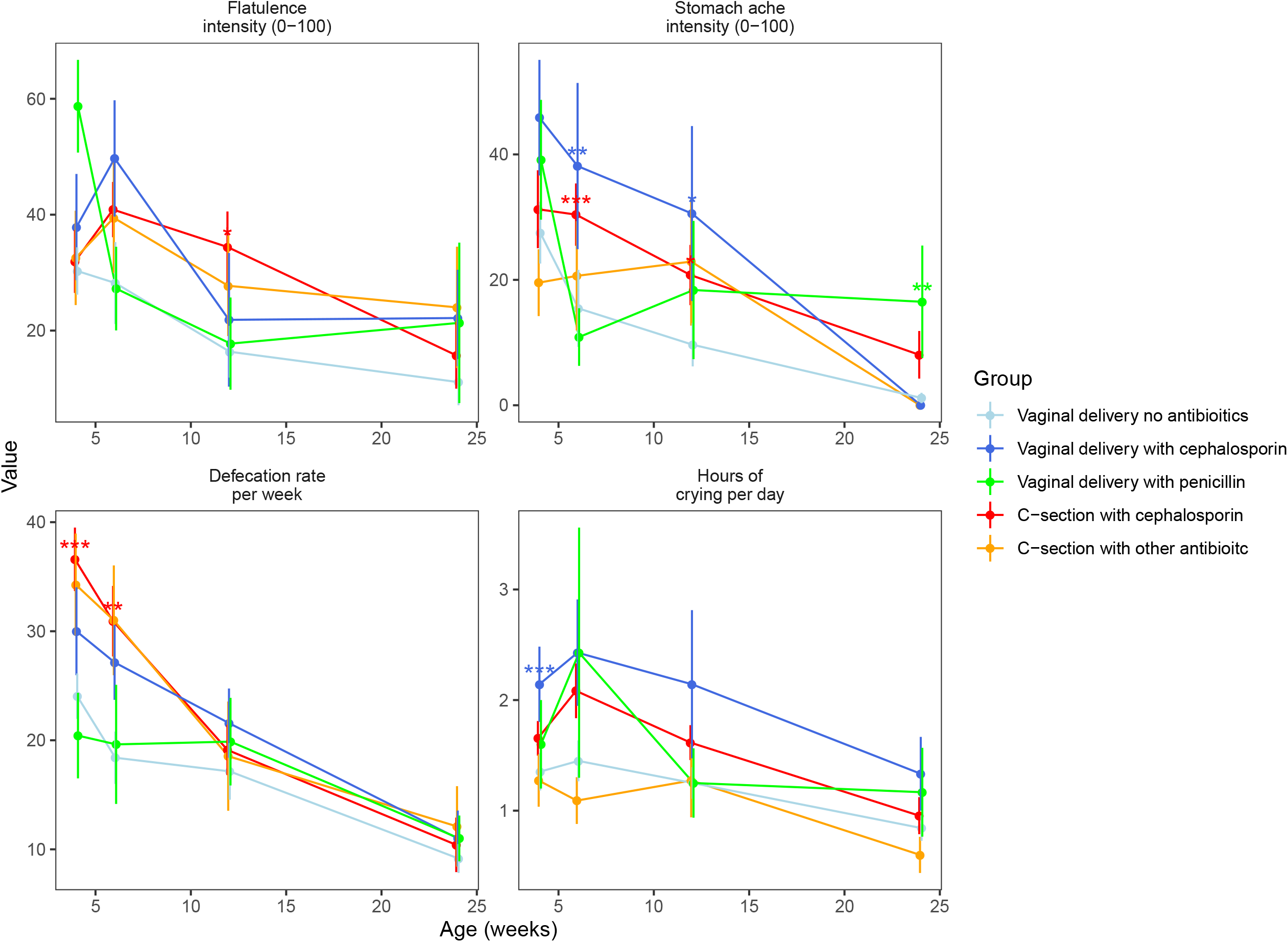
Parent-reported health outcomes at faecal sampling points from 1 to 6 by delivery group. *: *P*<0.05, **: *P*<0.01, ***: *P*<0.001

### Week 4

We assessed the hypothesis that the differences in the gut microbiota mediate the GI symptoms by performing a mediation analysis on the symptoms at time points that showed significant differences between the study groups (Figure 4) in infants with absolute data available (Supplementary Table 2). We analysed the association between antibiotic exposure and birth mode as separate variables with the health outcomes, compared the study groups in terms of the health outcomes, and identified bacterial taxa associated with both the health outcomes and the antibiotic exposure or birth mode. At 4 weeks, cephalosporin exposure (including both vaginal birth and CS) was significantly and positively associated with crying intensity (*P*=0.009), and when comparing groups, the VD-cep group had increased crying intensity in comparison to the reference group (*P*=0.009). A model using absolute bacterial abundances explained 24% of the variation in crying intensity (Figure 5a). The association of cephalosporin exposure with crying intensity appeared to be mediated by the low abundance of *Bacteroides* associated with cephalosporin exposure. Defecation rate was positively associated with cephalosporin exposure (*P*<0.0001) and CS (*P*<0.0001), and the CS-cep group had elevated defecation rate (*P*<0.0001) compared to the reference group. The bacterial composition explained 28% of the variation in defecation rate. The increased defecation rate in the CS-born and cephalosporin-exposed infants compared to the controls appeared attributable to the decreased abundances of *Bacteroides* and uncultured *Veillonellaceae* (Figure 5a).

**Figure 5.**
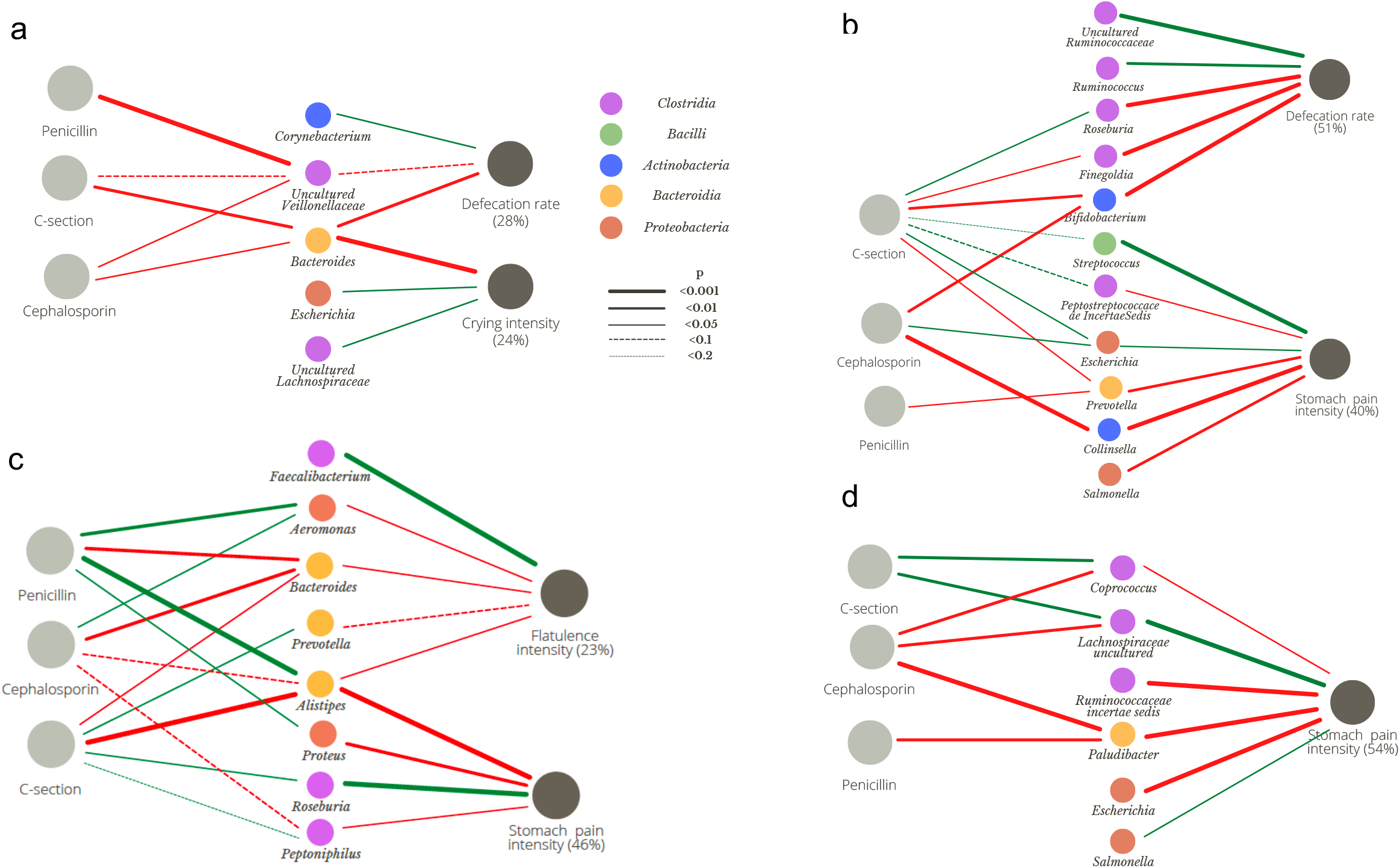
Associations between birth interventions, absolute abundances of bacteria, and GI symptoms at 4 weeks (a), 6 weeks (b), 12 weeks (c) and 24 weeks (d). The amount of variation explained by the bacteria shown is given as percentage. The models were selected based on Akaike information criterion (AIC). In some cases, the final model retained variables that were not statistically significant but contributed significantly to the model.

### Week 6

At 6 weeks, defecation rate was elevated in the CS-cep group (*P*=0.007) compared to the reference, and both CS birth (*P*=0.006) and cephalosporin exposure (*P*=0.01) were independently associated with increased defecation rate. The gut microbiota composition was associated with defecation rate, explaining 51% of the variation. The effects of CS birth and cephalosporin appeared to be mediated by the decreased abundances of *Bifidobacterium* and *Finegoldia* (Figure 5b). Stomach pain intensity was significantly elevated in the CS-cep (*P*=0.009) and VD-cep (*P*=0.01) groups compared to the reference. Cephalosporin exposure was significantly associated with stomach pain (*P*=0.005). The association was mediated by the reduced abundance of *Collinsella* and the increased abundances of *Escherichia* and *Streptococcus* in the CS-born and cep-exposed infants (Figure 5b). Gut microbiota composition explained 30% of the variation in stomach pain intensity.

### Week 12

At 12 weeks, stomach pain intensity was increased in the CS-cep group (*P*=0.04) and VD-cep group (*P*=0.02) compared to the reference. Cephalosporin exposure was significantly associated with the pain intensity (*P*=0.02). The gut microbiota explained 46% of the variation in stomach pain, the low absolute abundance of *Alistipes* and high abundance of *Roseburia* being linked with both CS birth and stomach pain (Figure 5c). Flatulence intensity was increased in the CS-cep group (*P*=0.02) compared to the reference, and CS birth (*P*=0.05) and cephalosporin exposure (*P*=0.02) were associated with increased flatulence intensity. The gut microbiota explained 23% of the variation in flatulence intensity at 12 weeks. The association of CS birth and cephalosporin and flatulence appeared mediated by the reduced abundance of *Bacteroides* and *Alistipes* (Figure 5c).

### Week 24

At 24 weeks, stomach pain intensity was associated with penicillin exposure (*P*=0.01) and increased in the VD-pen group (*P*=0.01) compared to the reference. Gut microbiota composition explained 54% of the variation in stomach pain intensity, with *Paludibacter* mediating the association with penicillin exposure (Figure 5d).

## Discussion

We set out to study how the temporal quantitative and compositional gut microbiota development is influenced by the mode of delivery versus exposure to delivery antibiotics, and whether these early exposures and associated differences in early microbiota composition relate to common gastrointestinal symptoms in infants. Whereas CS delivery had profound effects on the gut microbiota, we observed that maternal IP antibiotics compromised the colonisation of the newborn’s gut microbiota independent of the birth mode. Notably, the decreased absolute abundance of *Bifidobacterium* and increased abundance of Bacilli characterised both CS-deliveries and IP antibiotic exposed vaginally delivered infants, whereas reduction of *Bacteroides* was mainly seen after CS delivery. Moreover, we observed that the changes in gut microbiota taxa associated to CS birth and IP antibiotic exposure were further associated with increased GI symptoms.

This is one of the first studies using quantitative microbiota profiling^19, 36^ for community-wide absolute abundance estimates of infants’ indwelling gut bacteria and the first to address the associated GI health. The vast majority of existing NGS studies on human microbiota utilize only relative abundance. This has been recently shown to under- or overestimate temporal changes of the gut microbiota in preterm infants,^21^ especially concerning Proteobacteria such as *Escherichia coli* and *Klebsiella*. Several members within Proteobacteria, such as *E. coli*, possess flexible metabolic capacity^37^ and fast replication rate and therefore have high temporal variation in cell density. In the present study, we documented a decreasing developmental trajectory of the most abundant Proteobacterial order *Enterobacteriales* in relative abundance data, in line with earlier relative abundance studies.^38^ In contrast, the absolute abundance of *Enterobacteriales* increased over the first 6 months of life, peaking around the time of introducing complementary foods, in agreement with a previous study in term infants utilizing targeted qPCR.^39^ The developmental patterns of other dominant bacteria are largely in concordance with the previous qPCR-based study, showing a general upward trend in the first few months of life.^39^ Relative abundances also underestimate taxa with low 16S rRNA gene copy numbers, such as *Bifidobacteriales* (mean 3.5 copies^40^) compared to taxa with high copy numbers, such as *Clostridiales* (mean 5.5 copies^40^), observed also in this study.

CS birth has been suggested to cause microbial deprivation, a lack of microbial colonisation in early life. This was not the case in our study, as CS-born and antibiotic-exposed infants were colonised with comparable total bacterial loads per gram of faeces as the non-exposed vaginally born group. We observed a clear age-driven increase in the bacterial loads in all study groups using qPCR. Unfortunately, majority of the low bacterial biomass samples from the first week of life had insufficient amount of faecal sample to allow quantitative analyses. Previous studies have reported lower bacterial loads in CS born infants,^41^ and a faster age-driven increase in bacterial quantities in the stools of CS born infants.^42^ It is nevertheless possible that the transient difference in total microbial load occurring before 3 weeks of age was not detected in the present study.

Relative abundance studies have consistently documented an altered bacterial signature in gut microbiota of CS born infants,^1^ recently shown to be restored by maternal FMT.^43^ Most studies on infant microbiota in relation to birth mode have neglected the fact that practically all CS births involve IP antibiotics, making it impossible to isolate the effects of IP versus the birth route.^44^ Our results comparing the effect of birth mode under the exposure to the same antibiotic, cephalosporin, confirm that birth mode *per se* influences the infant gut microbiota. CS birth usually involves the administration of cephalosporin, while in VD, the most commonly used IP antibiotic is penicillin.^45^ It should be noted that the use of cephalosporin in VD typically indicates therapeutic rather than prophylactic use, and in our cohort, every other cep-VD mother received multiple doses compared to the single prophylactic dose used in CS. Hence, in clinical settings, labours with different IP antibiotics may also differ for other factors than the antibiotic exposure. A recent randomized trial documented a lack of clear differences in the gut microbiota of CS-born infants exposed versus unexposed to IP antibiotic.^45^ In conclusion, our study and earlier data indicate that the mode of birth is the major factor affecting neonatal colonisation, in line with CS eliminating the contact between the infant and maternal gut bacteria, preventing their transmission at birth.^46^ Infants born by elective CS, in the absence of labour and rupture of membranes, are believed to be the most deprived from the vertically transmitted microbes. A recent study however contradicted this hypothesis,^47^ which we corroborated by showing the negligible differences in the infant microbiota between the elective and urgent CS deliveries.

In contrast, in VD infants the IP antibiotic plays a major role. We showed that the impact of IP antibiotic exposure during vaginal birth on the gut microbiota was largely similar to the effect of CS delivery on the infant gut microbiota. However, we were able to identify several distinct effects of birth mode and antibiotic exposure. Namely, the early absolute abundances of *Bacteroidaceae, Coriobacteriaceae* and *Burkholderiaceae* were reduced by CS birth but not by IP antibiotics, while *Ruminococcaceae, Porphyromonadaceae, Rikenellaceae*, and *Pasteurellaceae* abundances were negatively affected by both CS birth and antibiotics. *Bifidobacteriaceae* was most strongly affected by the antibiotic exposure. These observations recapitulate and expand the results of a recent Japanese study^48^ and Canadian studies.^49^ With absolute data, we found the effect of IP antibiotics in VD infants to peak at 3 months with some differences persisting up to 12 months. Previous studies using relative microbiota data have reported the effect of birth-related antibiotics to resolve after 3 months,^47^ however, most of the studies do not extend over this age.^13^ A recent prospective study on 100 vaginally born infants reported the effect of IP antibiotics to overrule those of post-natal antibiotics on the infant microbiota, and persist until the at age of one year.^50^

Importantly, several bacterial taxa were increased in absolute abundance in the CS-born and in the antibiotic exposed infants. For example, IP antibiotic exposure in VD, and CS birth both resulted in the overgrowth of streptococci as well as staphylococci and enterococci. This is likely due to reduced colonization resistance resulting from the depletion of bifidobacteria, which have anti-streptococcal activity.^51^ In fact, the majority of bacterial taxa affected by IP antibiotics, except bifidobacteria, were increased in absolute abundance during the first month. This suggests that the major driver of microbial changes after exposure to IP antibiotic is not the antibiotic itself, but the antibiotic-induced reduction in bifidobacteria. Since the direct effects by the IP antibiotic on the infant are short (from hours to couple of days), the lasting effects likely reflect the consequences of the fundamental alteration in the ecosystem assembly.^52^ The pattern is somewhat different after CS birth, where diverse members of the microbiota, including *Bacteroides*, bifidobacteria and members of Clostridiales and Negativicutes, were likely directly reduced due to lower mother-infant microbiota transmission rates in CS birth.^7^

To our knowledge, our study is the first to evaluate the influence of birth mode and associated maternal antibiotic administration in parallel with the infant gut microbiota and functional GI symptoms. Previously, CS birth and postnatal antibiotics have been identified as risk factors for functional GI disorders in pre- and full-term infants.^53^ In our study, CS-born and VD IP antibiotic-exposed infants experienced more parent-reported abdominal discomfort and crying compared to VD infants with no antibiotics in the first month of life, while their general health and growth was comparable. According to the mediation analysis, the CS-associated reduction in the abundance of *Bacteroides* and its relatives was linked to increased defecation rates and crying intensity at 1 month, and increased flatulence and stomach pain at 3 months compared to the reference. The cephalosporin-associated decline in *Bifidobacterium* and *Collinsella* was associated with increased defecation rate and stomach pain at 6 weeks compared to the reference. Colic symptoms have previously been linked with the reduced relative abundance of bifidobacteria^54, 55^ and to an altered balance between H_2_-producing and -utilising taxa.^56^ Overall, the models based on absolute bacterial abundances explained 23-54% of the infants’ GI symptom variation, which is remarkable considering that the origin and underlying reasons of functional GI symptoms in infants often remain unclear.^57^ These results are preliminary and derived from a relatively small study, and hence need validation in future cohorts.

*Bacteroides* and bifidobacteria are the major human milk oligosaccharide (HMO) - degraders and thus keystone taxa in the breastfed infant gut,^58-60^ so their depletion will have profound impact on the metabolic capacity and processes in the infant gut.^61^ Reduced conversion of HMOs into short-chain fatty acids (SCFA) increases gut pH,^62, 63^ and alters trophic interactions and metabolic outputs of the gut microbiota. Such changes likely affect gut physiology and function and may favour bacteria that normally are less abundant in the infant gut, such as members of Clostridia, which in the present study were associated with increased defecation rates, flatulence, and stomach pain.

There is some uncertainty as to how long the microbiota differences persist after CS birth or IP antibiotic exposure, and how the persistence affects host physiology. Studies based on relative abundance data have shown a convergence to vaginally born controls during 6-12 months.^11, 64^ Long-term studies on the effects of IP antibiotic exposure have not been conducted. Our findings suggest that several birth-mode and IP antibiotic-related microbial signatures are still detectable at 12 months, although the major differences subside by 6 months. There may be individual differences regarding the normalisation of the gut microbiota after CS birth or antibiotic exposure, possibly related to diet, probiotic use, and other practices.^65, 66^ A recent Danish study showed that increased asthma risk was found in children born by CS only if their gut microbiota composition at 1 year of age still retained a CS-birth-related microbial signature,^9^ while animal experiments showcase that even a transient early microbiota disruption is sufficient to lead to persistent functional and even structural alterations in the immune system.^67^

Together, CS and antibiotic exposure at birth affect a substantial proportion of infants worldwide. Our data suggest that their effects on early colonisation of the gut microbiota may cause elevated levels of pain and discomfort in the infant during the first months of life, while a recent study found no association between IP antibiotics and childhood allergies.^18^ These studies are important primers for filling the knowledge gaps that underlie the lack of consensus for the choice between universal or risk-based screening for IP antibiotic use,^10^ and contribute to weighing the benefits and potential harms of antibiotic use in mothers and infants.

## Supporting information

Supplementary materials and results

## Data Availability

The sequencing data will be deposited upon acceptance of the article to European nucleotide Archive.

## Abbreviations

AIC: Akaike information criterion
ANOVA: Analysis of variance
Cep: Cephalosporin
CRP: C-reactive protein
CS: C-section, Caesarean section
GBS: Group B Streptococcus
GI: Gastrointestinal
HELMi: Finnish Health and Early Life Microbiota
HMO: Human milk oligosaccharide
IP: Intrapartum
NGS: Next generation sequencing
PCoA: Principal coordinate analysis
Pen: Penicillin
rRNA: Ribosomal ribonucleic acid
SCFA: Short-chain fatty acids
VAS: Visual analogue scale
VD: Vaginal delivery
qPCR: Quantitative polymerase chain reaction

## Acknowledgements

This research was supported by grants from Tekes 329/31/2015 (Health and the Early Life Microbiome (WMdV), Academy of Finland 1297765 (KK) and 1325103 (AS), Pediatric Research Foundation, Finland and Helsinki University Hospital Grant (KLK), The Paulo and Sohlberg Foundations (AS) and the Doctoral Program in Microbiology and Biotechnology, University of Helsinki, Finland funding (RJ). We thank Tinja Kanerva for processing the samples and Jessica Manngård for assistance with the questionnaire data analysis and Tytti Jaakkola, MD for recruiting some of the patients in the Jorvi cohort.

