## Supplementary materials and results for "Quantitative insights into effects of intrapartum antibiotics and birth mode on infant gut microbiota in relation to well-being during the first year of life"

### Supplementary Tables

Supplementary Table 1 Infant background variables summarised by study group.

| | | VD | VD-cep | VD-pen | CS-cep | CS-other |
|---|---|---|---|---|---|---|
| **Male (N=72, 50%)** | **Average birth weight (kg)** | 3.56 SD=0.47 N=26 | 3.50 SD=0.46 N=8 | 3.36 SD=0.50 N=12 | 3.62 SD=0.42 N=17 | 3.71 SD=0.61 N=9 |
| | **Average weight at 1 year (kg)** | 9.54 SD=0.88 N=24 | 10.7 SD=1.63 N=4 | 9.63 SD=0.97 N=8 | 9.74 SD=0.69 N=15 | 10.1 SD=0.72 N=9 |
| **Female (N=72, 50%)** | **Average birth weight (kg)** | 3.49 SD=0.39 N=32 | 3.53 SD=0.24 N=5 | 3.37 SD=0.45 N=13 | 3.42 SD=0.42 N=17 | 3.64 SD=0.56 N=5 |
| | **Average weight at 1 year (kg)** | 9.39 SD=0.97 N=24 | 9.42 SD=0.87 N=5 | 9.08 SD=0.94 N=10 | 9.32 SD=1.4 N=13 | 9.91 SD=1.6 N=5 |
| **Breast-feeding** | **Exclusive at 3 months** | 80.5% | 91.7% | 54.6% | 77.4% | 76.9% |
| | **Any 3 months** | 95.1% | 100% | 100% | 93.6% | 100% |
| | **Any 6 months** | 91.7% | 91.7% | 100% | 90.6% | 92.3% |
| | **Any 9 months** | 93.8% | 85.7% | 88.9% | 80.0% | 80.0% |
| | **Any 12 months** | 72.2% | 62.5% | 60.0% | 75.0% | 66.7% |
| **Solid foods** | **Age of introduction (months)** | 4.63 SD=0.94 | 4.69 SD=0.85 | 4.24 SD=0.52 | 4.71 SD=0.87 | 4.79 SD=0.89 |
| **Pro-biotic intake** | **During 1st year** | 94.8% | 53.5% | 72.0% | 85.3% | 57.1% |

VD = vaginal delivery, CS = Caesarean section, cep = cephalosporin, pen = penicillin, SD = standard deviation

Supplementary Table 2 Group sizes for gastrointestinal symptom mediation analysis.

| Age (weeks) | VD | VD-cep | VD-pen | CS-cep | CS-other | Total |
|---|---|---|---|---|---|---|

| | | | | | | |
|---|---|---|---|---|---|---|
| 4 | 21 | 7 | 7 | 27 | 11 | 73 |
| 6 | 20 | 7 | 8 | 25 | 11 | 71 |
| 12 | 20 | 7 | 8 | 26 | 11 | 72 |
| 24 | 19 | 6 | 6 | 22 | 10 | 63 |
| 36 | 16 | 4 | 6 | 20 | 7 | 53 |

VD = vaginal delivery, CS = Caesarean section, cep = cephalosporin, pen = penicillin

### Supplementary Figures

Supplementary Figure 1 Differences in bacterial families between the two C-section (CS) types.

Supplementary Figure 2 Logarithmically-transformed total bacterial counts at different age grouped by delivery group. No significant difference (*P*>0.5) was found between delivery group at any given age.

Supplementary Figure 3 Fold changes calculated using absolute abundances of bacterial genera with at least one group significantly differing from the reference group (VD) in the taxon-wise comparisons between all delivery groups. VD: vaginal delivery without antibiotic, VD-cep: vaginal delivery with cephalosporin, VD-pen: vaginal with penicillin, CS-cep: C-section with cephalosporin, CS-other: C-section with any other antibiotic; FC: fold change (cut-off at 5 and 0.001 for clarity); *: P<0.05, **: P<0.01, ***: P<0.001, FDR<0.1.

Supplementary Figure 4 Fold changes calculated using absolute abundances of bacterial families with at least one group significantly differing from the VD-cep group in the taxon-wise comparisons. VD-cep: vaginal delivery with cephalosporin, VD-pen: vaginal with penicillin, CS-cep: C-section with cephalosporin; FC: fold change (cut-off at 5 and 0.001 for clarity); *: P<0.05, **: P<0.01, ***: P<0.001, FDR<0.1.

Supplementary Figure 5 Fold changes calculated using absolute abundances of bacterial genera with at least one group significantly differing from the VD-cep group in the taxon-wise comparisons. VD-cep: vaginal delivery with cephalosporin, VD-pen: vaginal with penicillin, CS-cep: C-section with cephalosporin; FC: fold change (cut-off at 5 and 0.001 for clarity); *: P<0.05, **: P<0.01, ***: P<0.001, FDR<0.1.

### Supplementary methods

#### Microbiota profiling

Faecal DNA was extracted from *ca.* 125 mg of fecal material that was suspended in 1 ml of sterile ice-cold PBS, and 175 µl of fecal suspension was combined with 235 µl of RBB lysis buffer (500 mM NaCl, 50 mM Tris-HCl (pH 8.0), 50 mM EDTA, 4% SDS) in a bead-beating tube from the Ambion Magmax™ Total Nucleic Acid Isolation Kit (Life Technologies, Carlsbad, CA, USA). After repeated bead-beating, 200 µl of the supernatant was used for DNA extraction with a KingFisher™ Flex automated purification system (ThermoFisher Scientific, Waltham, MA, USA) using a MagMAX™ Pathogen High Vol. DNA was quantified using Quanti-iT™ Pico Green dsDNA Assay (Invitrogen, San Diego, CA, USA).

Due to low DNA and/or read yields, for 161 samples the V3-V4 library preparation protocol was modified with one or more of the following modifications: additional DNA concentration with ethanol precipitation, 2-step index PCR, increased input of template DNA (5 vs 1 ng/rx), more PCR cycles (45 vs 27).

#### Statistical analysis

The effects of birth mode and IP antibiotic on the abundances of the bacterial taxa were analysed using negative binomial, Poisson or quasi-Poisson models, depending on data distribution and model fit, using the VD group without antibiotics as the reference group. If the fitted model failed to fulfil model assumptions (primarily heteroscedasticity of the residuals), generalised least squares models were used. Only genera observed in >30% of the samples were analysed individually. Results with FDR adjusted *P*-values <0.1 were considered significant. All models were adjusted for infant probiotic intake at the time of sampling (none, *Lactobacillus* spp., *Bifidobacterium* spp. or both, or *Saccharomyces* spp.), feeding type, including breastfeeding status (none, partial, exclusive) and weeks since the introduction of solids, and the sample treatment history, including possible PCR modifications and sequencing run ID. The health outcome variables were compared between the birth groups at each faecal sampling until 9 months using a negative binomial model adjusting for feeding type and probiotic use.

### Supplementary results

We tested whether the type of CS (elective versus urgent) was associated with the gut microbiota composition in infants before pooling the different CS types. The relative abundance of *Bacteroides* was reduced in elective compared to urgent CS at week 1, 3 and 6 (3.6-8.9-fold difference, $P<0.005$), and 6 months (5.1-fold difference, $P=0.02$). The difference in absolute abundance was not significant. *Bifidobacterium* was significantly reduced at 6 months in elective CS vs urgent? in both relative abundance (9.8-fold difference, $P=0.002$) and absolute abundance (5.7-fold difference, $P=0.03$). Although there were subtle differences between the microbiota after urgent and elective CS (Supplementary Figure 1), the differences between the microbiota of CS versus vaginally-delivered infants (independent of IP antibiotics) remained large even after pooling, justifying the grouping of CS types (Table 1).

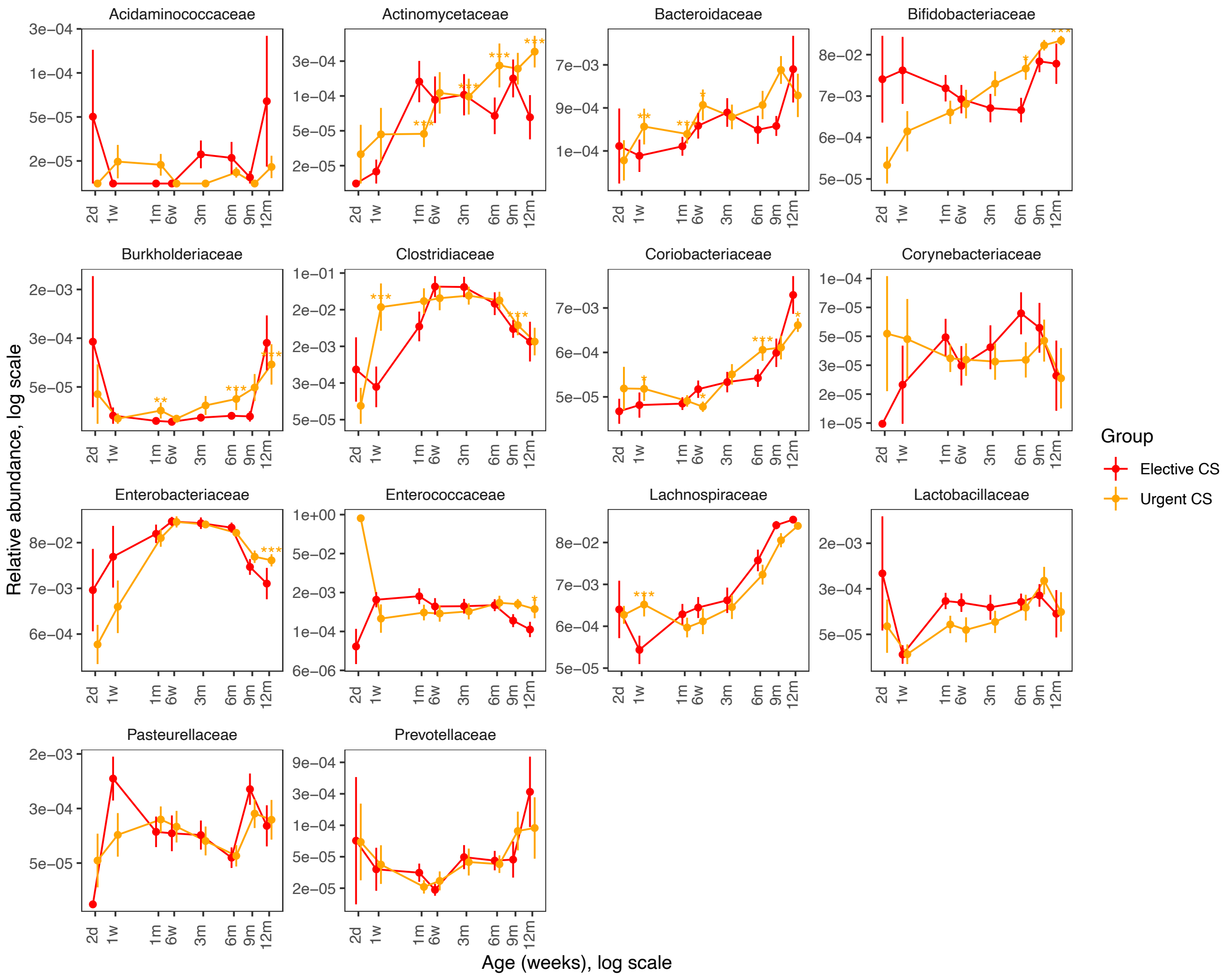


### Supplementary_figure1

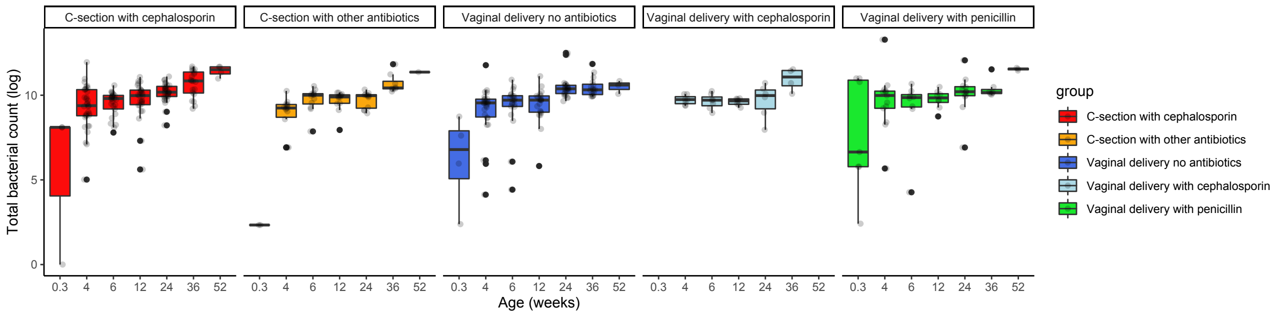


### Supplementary_figure2

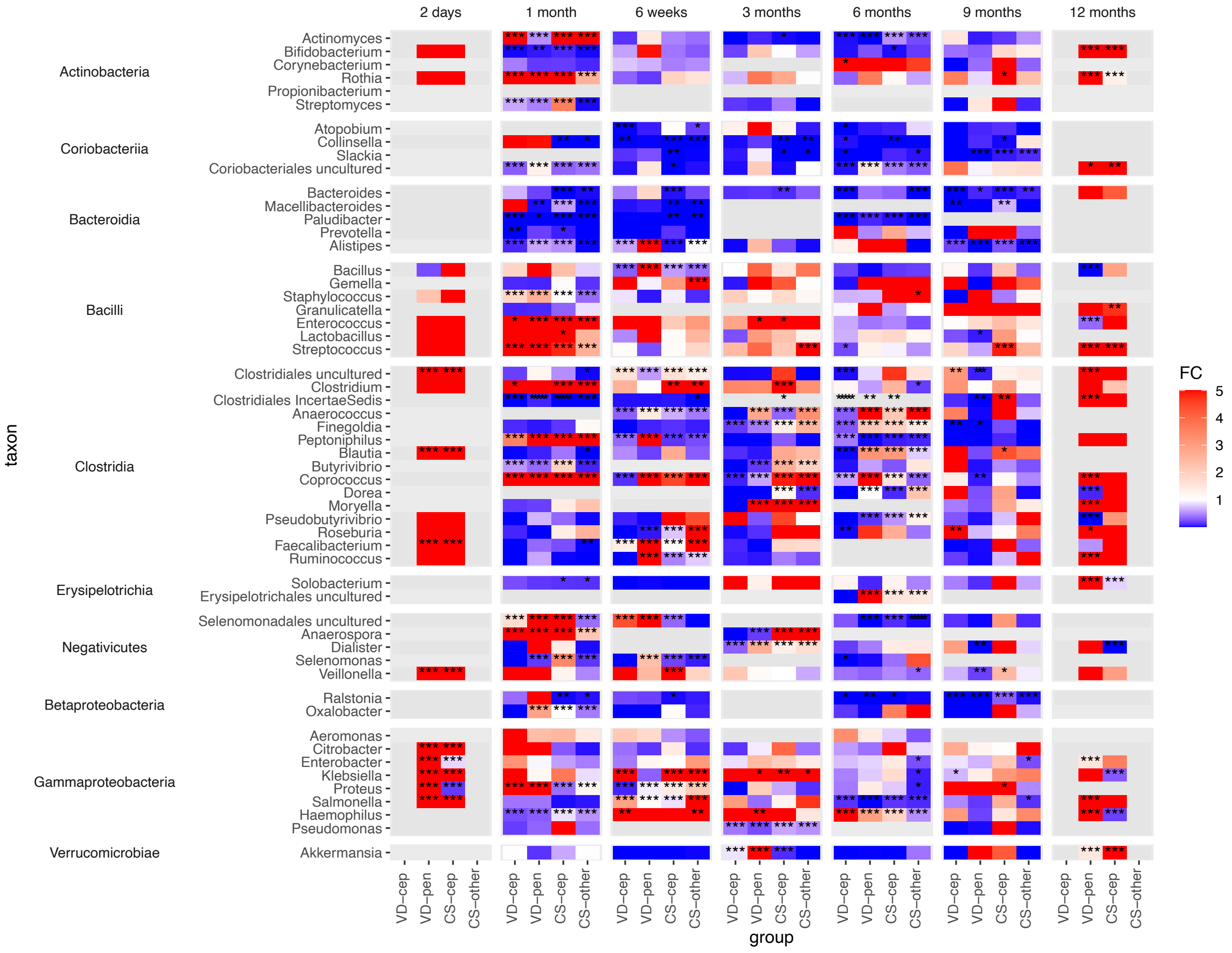


**Supplementary_figure3**

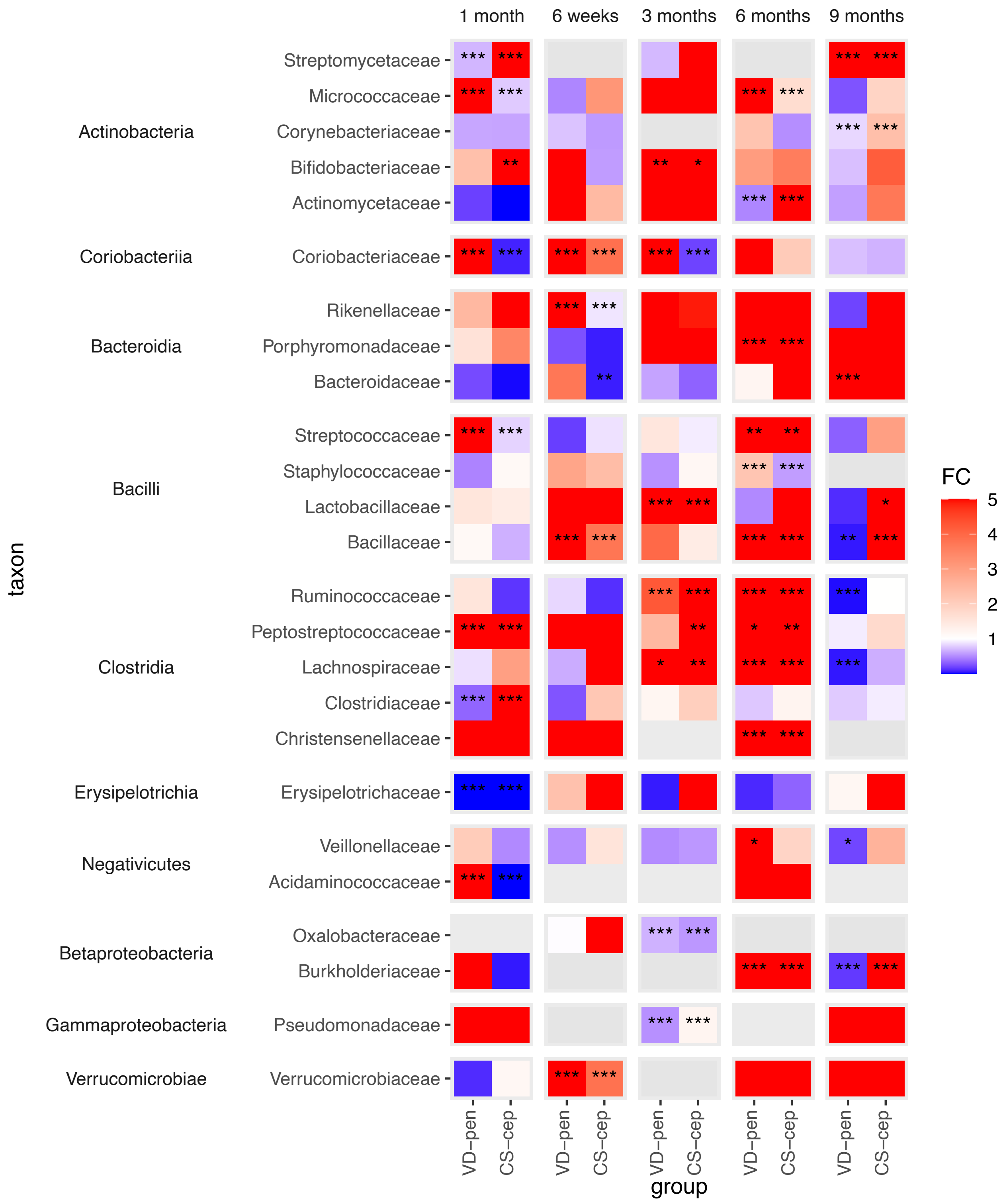


**Supplementary_figure4**

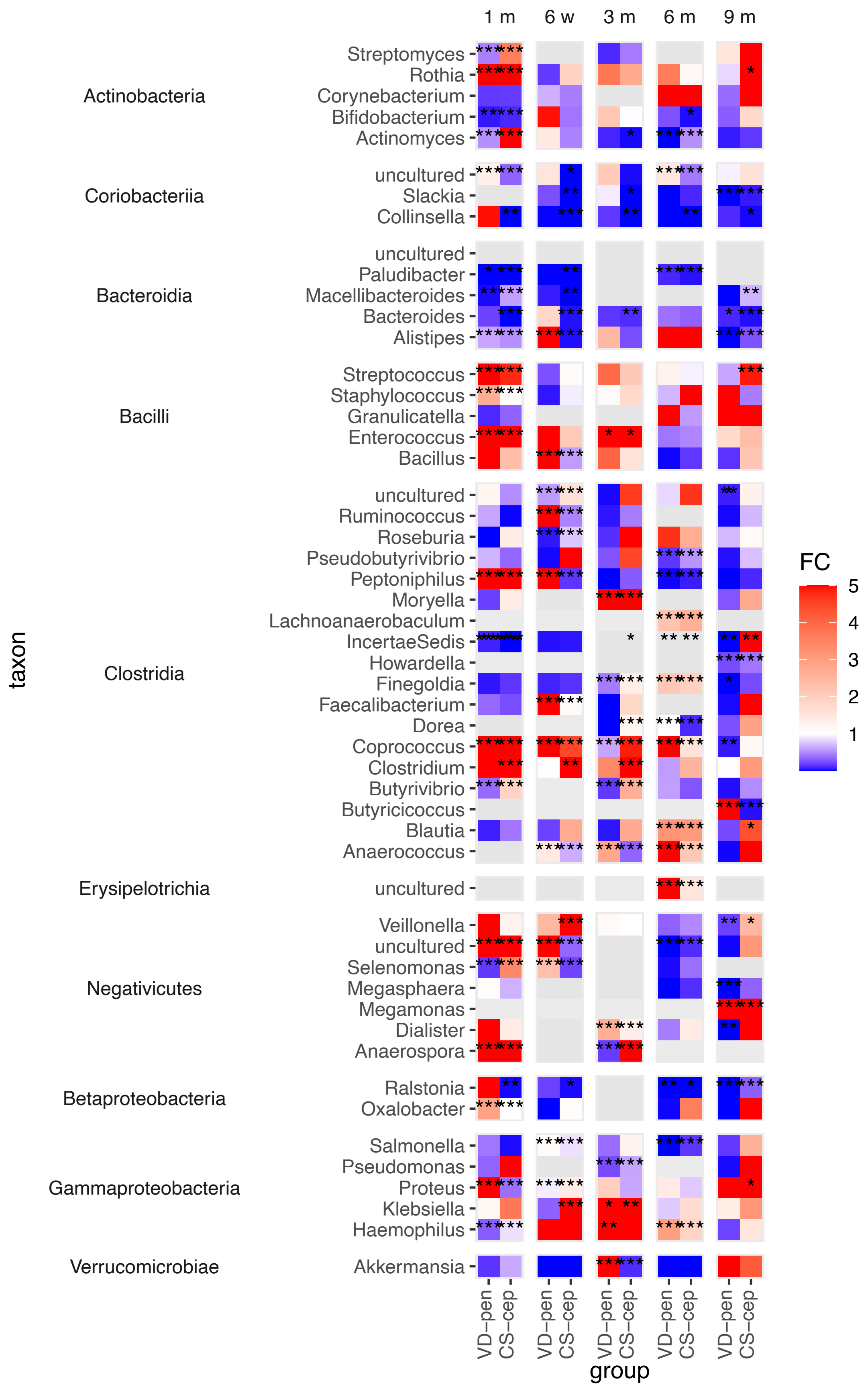


**Supplementary_figure5**
